# Performance evaluation of the Simtomax^®^ CoronaCheck rapid diagnostic test

**DOI:** 10.1101/2020.10.28.20219667

**Authors:** P. J. Ducrest, A. Freymond, J.-M. Segura

## Abstract

The aim of this study was to evaluate the diagnostic performance of Simtomax^®^ CoronaCheck, a serology rapid diagnostic test (RDT) for the detection of IgG and IgM against SARS-CoV-2. 48 plasma samples positive for SARS-CoV-2 based on RT-PCR and 98 negative control samples were studied. Diagnostic performance of the IgG/IgM RDT was assessed against RT-PCR and the electro-chemiluminescence immunoassay (ECLIA) Elecsys^®^ Anti-SARS-CoV-2 total Ig. Overall, the RDT sensitivity was 92% (95% confidence interval [95%CI]: 79-97), specificity 97% (95% CI: 91-99%), PPV 94% (95% CI: 81-98) and the NPV 96% (95% CI: 89-99). When considering only samples collected ≥ 15 days post-symptoms (DPS), the sensitivity increased to 98% (95%CI: 86-100) and the specificity was 97% (95% CI: 91-99%). Two samples with 180 DPS were still positive for IgG. Globally, this IgG/IgM RDT displayed a high diagnostic accuracy for SARS-CoV-2 IgG/IgM detection in plasma samples in high COVID-19 prevalence settings. It could be effectively used, in absence of facilities for routine diagnostic serology, for samples with a DPS between 15 and 180 days.

**Highlights:**

– The rapid diagnostic test Simtomax CoronaCheck displays a high sensitivity of 98% and a high specificity of 97% for SARS-CoV-2 IgG/IgM detection in plasma samples after 15 days post-symptoms.
– The rapid diagnostic test Simtomax CoronaCheck can detect SARS-CoV-2 antibodies in plasma up to 180 days after symptom onset.
– The rapid diagnostic test Simtomax CoronaCheck could be effectively used as an alternative to serological analysis using laboratory facilities.

## Introduction

SARS-CoV-2 is the etiological agent of a severe pneumonia first reported in Wuhan (Hubei, China), called 2019 Coronavirus Disease (COVID-19). Protein sequence analysis of seven proteins showed that the virus is related to the Severe Acute Respiratory Syndrome Coronavirus (SARS-CoV) and the Middle East Respiratory Syndrome Coronavirus (MERS-CoV) with similar epidemiology (Kannan et al. 2020; Zhou et al. 2020). Currently, the detection method for SARS-CoV-2 is based on viral RNA detection using Reverse-Transcription PCR (RT-PCR) but other tests such as chest computed tomography (CT) imaging or antigen/antibody testing can also be used (Li et al. 2020). Testing of specific antibodies against SARS-CoV-2 in patient blood offers rapid, simple, highly sensitive diagnosis of COVID-19. Moreover, serological analysis may be applied to detect past exposure to the virus and possibly if a patient has developed immunity against the virus. It is widely accepted that IgM provide the first line of defense during viral infections, prior to the generation of adaptive, high affinity IgG responses that are important for long term immunity and immunological memory (Li et al. 2020b). Several authors analyzed the antibody kinetics in COVID-19 patients. Zhao et al. (2020) showed that among 173 patients, the seroconversion sequentially appeared for total antibody, IgM and then IgG, with a median time of 11, 12 and 14 days after symptom onset. The majority of antibodies are produced against the most abundant protein of the virus, which is the Nucleocapsid protein (NP). Therefore, serological tests that detect antibodies to NP should be the most sensitive. In addition, because the receptor-binding domain of the Spike protein (RBD-S) is the host attachment protein, antibodies against RBD-S should be very specific. Therefore, according to some authors, using one or both antigens should result in high sensitivity and specificity (Sethuraman et al. 2020). The rapid diagnostic test (RDT) Simtomax® CoronaCheck developed by Augurix SA (Switzerland) uses both antigens as described in To et al. (2020). The gold nanoparticles used for detection are conjugated with SARS-CoV-2 Receptor Binding Domain and Nucleocapsid protein with the aim to specifically bind IgM and/or IgG in COVID-19 positive samples. A recent clinical evaluation of Augurix RDT at the University Hospitals of Geneva and Lausanne (Switzerland) demonstrated high diagnostic accuracy for IgG in whole-blood, plasma and sera samples (Andrey et al. 2020; Coste et al. 2020).

The aim of this study was to evaluate the performance of this RDT in the case of plasma samples in a high COVID-19 prevalence setting using as reference methods RT-PCR and an Electro-chemiluminescence immunoassay (ECLIA) Elecsys® Anti-SARS-CoV-2 total Ig (Roche, Switzerland).

## Materials & Methods

### Study population and blood sample collection

48 anonymized leftovers of diagnostic plasma-EDTA specimen, supplied by INO Specimens BioBank, ISB (Clermont-Ferrand, France), were used for this method evaluation (Fig. 1). Only laboratory-based information was used in this study. All 48 samples were positive for SARS-CoV-2 based on RT-PCR (Information provided by INO Specimens based on analysis using the BD SARS-CoV-2 reagent kit for the BD Max system; Becton Dickinson and Co, US. All RT-PCR tests were done on nasopharyngeal secretions. The median CT value was 25.6 (IQR 20.05-29.15)) and had been collected at days post symptom (DPS) of at least 10 days. 23 out of 48 RT-PCR positive samples were additionally tested for total immunoglobulin against COVID-19 with an electro-chemiluminescence immunoassay (ECLIA) at INO Specimens BioBank (Figure 1). As negative controls, anonymized unmatched control plasma samples (n=98), supplied by AbBaltis (Kent, UK) with a collection date before 2018, were used (Fig. 1).

**Figure 1:**
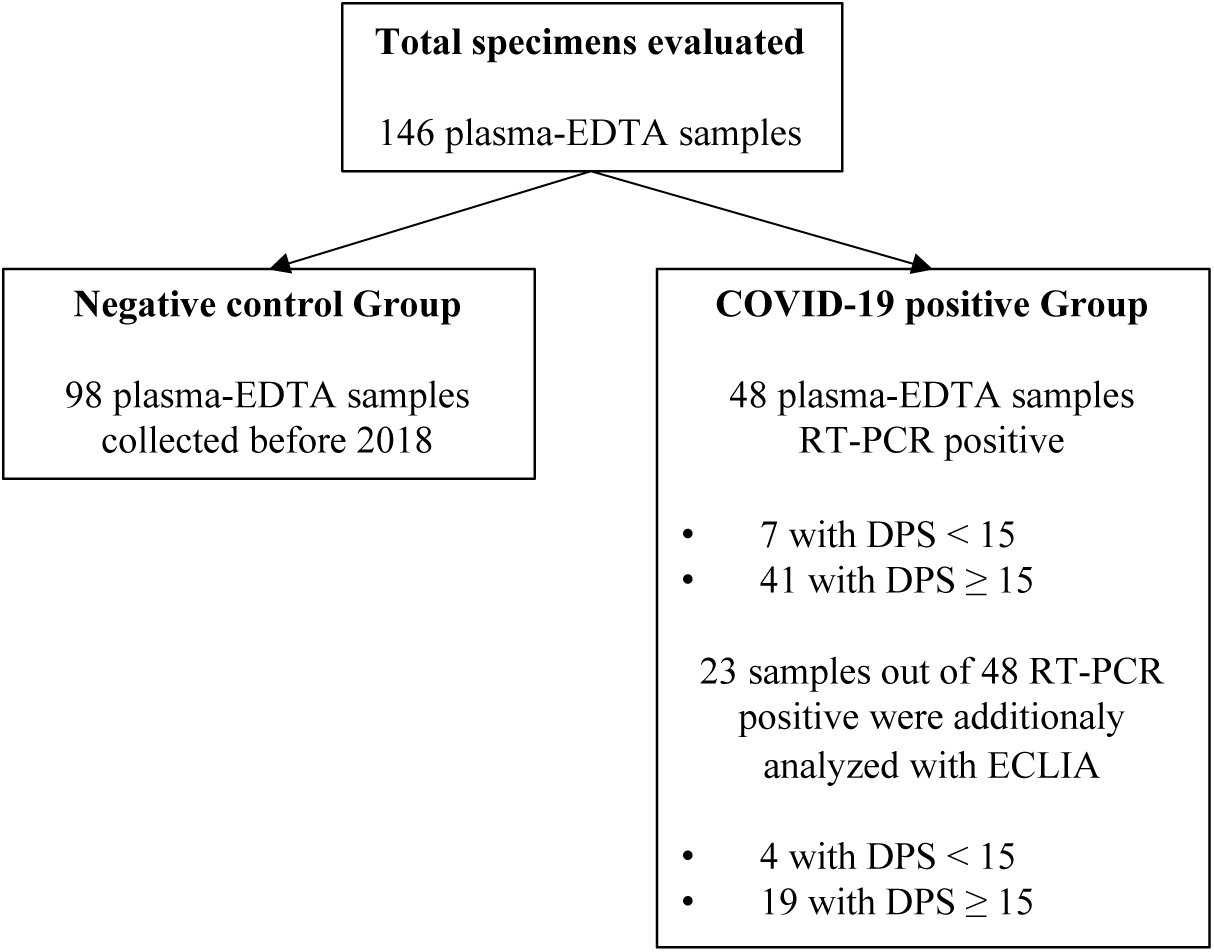
Description of the plasma samples used in this study.

### Augurix IgM/IgG immunochromatographic rapid test

Commercial CE-labeled SARS-CoV-2 IgM/IgG RDT were from Augurix (Switzerland). These tests can be used either with capillary blood, whole blood, plasma or serum. One IgM/IgG rapid test per sample was used. Following manufacturers’ instructions, 10 μL of plasma were applied for each sample. IgG and IgM responses were read after 15 minutes following manufacturer’s instructions, blinded to the reference method results. The tests were considered COVID-19 positive if either the IgM line or the IgG line or both lines were positive.

### Electro-chemiluminescence immunoassay Elecsys® Anti-SARS-CoV-2 total Ig

ECLIA experiments were performed at INO Specimens Biobank. Elecsys® Anti-SARS-CoV-2 total Ig (Roche, Switzerland) makes use of the nucleocapsid protein of SARS-CoV-2 as antigen. Total Ig was analyzed according to manufacturer’s instructions. The assays were run on a Cobas e 601 (Roche, Switzerland) according to manufacturer’s protocol. Positivity was defined by the manufacturer as a cut-off index (COI) ≥ 1.0.

### Study end points

The primary end point was to assess the accuracy of IgG/IgM detection in plasma using Augurix IgM/IgG RDT against the RT-PCR reference method, within a cohort of 48 RT-PCR confirmed COVID-19, with day post-symptoms (DPS) of ≥ 10 days, and 98 control plasma samples. The secondary end point was to assess Augurix IgG/IgM RDT performance against Electro-chemiluminescence immunoassay (ECLIA), Elecsys® Anti-SARS-CoV-2 total Ig (Roche, Switzerland).

### Statistics

Vassarstats online tool (www.vassarstats.net) was used to calculate sensitivity (SE), specificity (SP), positive and negative predictive values (PPV, NPV), 95% confidence intervals, median, and Interquartile range (IQR). Significance (p-values) were calculated using a Mann-Whitney U test. Statistical significance was defined as p < 0.05.

## Results

### Baseline characteristics

The demographic characteristics of patients’ samples was as follows: the 48 COVID-19-positive samples were from patients older (median = 49 years old, IQR 33-58.75) compared to the healthy patients (n=98) (median = 34.5 years old, IQR 18-44.75; p < 0.05). The proportion of females was 56% (n=20) and 51% (n=50) in the COVID-19 positive and in the healthy control group, respectively. Among the COVID-19 samples, the median delay between symptom onset and sampling was 21 days (IQR 16-32 days), but not less than 10 days. The longest DPS (one single sample) was 180 days. A detailed description of all the available information on the samples and the results of the various tests is provided in the supplementary information.

### Specificity of IgG/IgM RDT on the negative control group

The diagnostic specificity of the IgG/IgM RDT was assessed on the COVID-19 negative control group with a sampling date before 2018 (n=98). The results are shown in Table 1 and 2. IgG/IgM RDT results were negative in 95 of the 98 samples, i.e. in 96.9% of the cases (95%CI: 91-100%). Three discordant results showed a positive IgM line (false positives). The specificity (SP) of the IgG/IgM RDT was therefore 97% (95% CI: 91-99%).

**Table 1:** Diagnostic performance of Simtomax CoronaCheck RDT compared to RT-PCR as reference method for different sample groups.

|  | SE %<br>(95%CI) | SP %<br>(95%CI) | PPV %<br>(95%CI) | NPV %<br>(95%CI) |
| --- | --- | --- | --- | --- |
| **All samples (n=146); Prevalence = $48 / 146 = 33\%$** |  |  |  |  |
| **IgG/IgM RDT vs RT-PCR** | 92 (79-97) | 97 (91-99) | 94 (81-98) | 96 (89-99) |
| **Samples with DPS < 15 and negative controls (n=105); Prevalence = $7 / 105 = 6.7\%$** |  |  |  |  |
| **IgG/IgM RDT vs RT-PCR** | 57 (20-88) | 97 (91-99) | 57 (20-88) | 97 (91-99) |
| **Samples with DPS $\geq 15$ DPS and negative controls (n=139); Prevalence = $41 / 139 = 29\%$** |  |  |  |  |
| **IgG/IgM RDT vs RT-PCR** | 98 (86-100) | 97 (91-99) | 93 (80-98) | 99 (94-100) |

RDT: Simtomax CoronaCheck rapid diagnostic test; SE: sensibility; SP: specificity; PPV: positive predictive value; NPV: negative predictive value; ECLIA: Electro-chemiluminescence immunoassay

**Table 2:** Diagnostic performance of Simtomax CoronaCheck RDT compared to ECLIA total Ig as reference method for different sample groups.

|  | **SE %**<br>**(95%CI)** | **SP %**<br>**(95%CI)** | **PPV %**<br>**(95%CI)** | **NPV %**<br>**(95%CI)** |
| --- | --- | --- | --- | --- |
| **All samples (n=121); Prevalence = 23 / 121 = 19%** |  |  |  |  |
| **IgG/IgM RDT vs ECLIA** | 91 (70-98) | 97 (91-99) | 88 (67-97) | 98 (92-100) |
| **Samples with DPS < 15 and negative controls (n=102); Prevalence = 4 / 102 = 3.9 %** |  |  |  |  |
| **IgG/IgM RDT vs ECLIA** | 50 (9-91) | 97 (91-99) | 40 (7-83) | 98 (92-100) |
| **Samples with DPS ≥ 15 DPS and negative controls (n=117); Prevalence = 19 / 117 = 16 %** |  |  |  |  |
| **IgG/IgM RDT vs ECLIA** | 100 (79-100) | 97 (91-99) | 86 (64-96) | 100 (95-100) |

### Sensitivity of IgG/IgM RDT on RT-PCR confirmed COVID-19 samples

IgG/IgM RDT diagnostic sensitivity was assessed on the 48 samples positive for COVID-19 based on RT-PCR. The results are shown in Table 1. Both methods revealed similar results in 44 of 48 samples, i.e. 91.7% (95%CI: 79-97%). Four discordant results showed a negative result with the IgG/IgM RDT both for IgG and IgM (false negatives). The resulting sensitivity (SE) of the IgG/IgM RDT was 92% (95% CI: 79-97), while the positive predictive value (PPV; using a prevalence of 48 / 146 = 33%) was 94% (95% CI: 81-98) and the negative predictive value (NPV) 96% (95% CI: 89-99).

As shown in Table 1, most false-negative results (3 out of 4) exhibited a DPS between 10 and 15. Only one false-negative result was observed with a DPS ≥ 15. When taking into account exclusively the results in the ≥ 15 DPS group, the IgG/IgM RDT sensitivity (SE) was 98% (95% CI: 86-100), while the PPV (using a prevalence of 41 / 139 = 29%) was 93% (95% CI: 80-98) and the NPV 99% (95% CI: 94-100). It is remarkable that two samples with days post symptoms (DPS) of respectively 170 and 180 days were still positive for IgG (Sup. Information).

### Sensitivity of IgG/IgM RDT with an electro-chemiluminescence immunoassay as reference method

The accuracy of the IgG/IgM RDT was also assessed using an electro-chemiluminescence immunoassay (ECLIA), Elecsys® Anti-SARS-CoV-2 total Ig, as reference method. ECLIA was conducted on a random selection of 23 out of the 48 COVID-19 positive samples based on RT-PCR. The median cut-off index (COI) was 63.4 (IQR 5-85 COI). The COI was not significant higher in the ≥ 15 DPS group (76.9; IQR 5.8-86) compared to the < 15 DPS group (26.3; IQR 1-49.5; p=0.384). This might be at least partly attributed to the small sample size of the < 15 DPS group (n=4). The results obtained using the IgG/IgM RDT on the 23 samples are shown in Table 2. Both methods yielded similar results in 21 of the 23 samples, i.e. in 91.3% of the cases (95%CI: 70-98%). Two discordant results showed an IgG/IgM RDT negative result, both for IgG and IgM, while were positive by ECLIA (false negatives). This resulted in an IgG/IgM RDT sensitivity (SE) of 91% (95% CI: 70-98), while the PPV (using a prevalence of 23 / 121 = 19%) was 88% (95% CI: 67-97) and the NPV 98% (95% CI: 92-100) compared to ECLIA. The two false-negative results exhibited a DPS between 10 and 15. When using exclusively the results in the ≥ 15 DPS group, there was a complete agreement between the results of the IgG/IgM RDT and ECLIA. In this case, the IgG/IgM RDT sensitivity (SE) was therefore 100% (95% CI: 79-100), while the PPV was 86% (95% CI: 64-96) and the NPV 100% (95% CI: 95-100).

## Discussion

The main finding of this evaluation study, using an unmatched case-control design including 67.1% of negative control samples, is that the diagnostic accuracy of IgG/IgM Augurix RDT on plasma samples when compared to RT-PCR confirmed cases displayed a SE of 92%, a SP of 97%, a PPV of 94% and a NPV of 96%. When compared to ECLIA positive samples, the diagnostic accuracy of IgG/IgM Augurix RDT displayed a SE of 91%, a SP of 97%, a PPV of 88% and a NPV of 98%. The sensitivity (SE) of IgG/IgM Augurix RDT is not significantly different for both reference methods (p=0.756) indicating a good correlation. The diagnostic accuracy of the RDT further increased when analyses were performed exclusively on samples collected after 15 DPS and exhibited excellent sensitivity (98%) and NPV (99%). For samples collected within a DPS < 15 days, the diagnostic performance was clearly poorer with 57%, 97%, 57% and 97% for SE, SP, PPV and NPV, respectively.

It is interesting to notice that the two false-negative results obtained with Augurix RDT corresponded to borderline samples in ECLIA (Cut-of index of 1.43 and 1.19) with a DPS between 10 and 15 days. The ECLIA manufacturer defines positive samples when the cut-off index (COI) value is ≥ 1.0. This finding indicates that the analytical sensitivity of Augurix RDT is lower than the ECLIA Elecsys® Anti-SARS-CoV-2 total Ig (Roche, Switzerland) and that insufficient analytical sensitivity might at least partly explain the poor performance of the RDT test at DPS < 15 days.

For samples with DPS > 15, there is a perfect agreement between ECLIA and Augurix RDT, which can be explained by the fact that both assays target the immune response against the full-length N protein of SARS-CoV-2. The overall good specificity of the Augurix RDT might be offered by the fact that it also targets the immune response against the Receptor binding domain (RBD) of S protein,

The overall performance, in particular for DPS > 15, indicates that Augurix RDT could be fit for purpose in clinical settings where a high prevalence of COVID-19 prevails, especially in situations where ECLIA is not available or cannot be reliably used. Diagnostic performance in low prevalence populations still needs to be experimentally determined and it would be interesting to validate the results on larger populations. The results of our study on plasma samples are similar with those recently published on whole blood and plasma samples, which indicated a sensitivity of Augurix RDT between 93% and 100% for samples with a DPS of >15 days. By contrast, our results differ from prior studies using Augurix and other RDTs that observed a lower sensitivity of 56.4% for a DPS of > 21 days (Rudolf et al. 2020). A first difference between the present study and the one of Rudolf et al. (2020) is that positive samples were assessed here when either the IgM or the IgG band was positive, while in Rudolf et al, the results were separated for the two lines, i.e. they assessed sensitivity for either the IgM or the IgG line. A second possible partial explanation for the discrepancy is that the study by Rudolf et al included a large panel of samples corresponding to varying severities of the symptoms. As this information is not available for the plasma samples analyzed in our study, it is possible that they were collected from patients showing more severe forms of COVID-19. On the other hand, the antibody titers in the plasma samples as determined by ECLIA spanned a large range of concentration, which points towards the fact that the samples in our study corresponded to various severities of COVID-19. A more comprehensive study on a larger panel of well-defined sample might clarify these discrepancies. Concerning the specificity of the test, the excellent performance observed in this study is in line with the results of preceding publications (Andrey et al. 2020; Coste et al. 2020). Three control blood sample turned out to be IgM positive by RDT. These samples were collected before 2018 and it seems that a cross-reaction with the IgM line occurred leading to a false positive result. But overall, Augurix RDT might be a suitable choice in situations where a high PPV is instrumental.

The second notable finding of this study lies in the fact that IgG seropositivity is still present 180 days after symptom onset in spite of normal antibody decline (Long et al. 2020; Seow et al. 2020). To our knowledge, it is the first time that SARS-CoV-2 IgG seropositivity is demonstrated with RDT 180 DPS. This study indicates also that a certain titer of SARS-CoV-2 IgG is present constantly with a concentration sufficient to be detectable with RDT, from 15 days to at least 180 days post symptoms. This finding applies to the Augurix RDT and cannot be generalized to other RDTs currently available. It would be interesting to quantitatively determine the level of IgG/IgM 180 days post symptoms onset to confirm this finding obtained with a qualitative assay.

In addition, the test provided clear results without indeterminate or invalid measurements. There are however several limitations to this study. First, we present here the results of a method evaluation study and not a seroprevalence study. Therefore, the PPV obtained here (based on a 32.9% proportion of cases defined as laboratory confirmed SARS-CoV-2 by RT-PCR) will be lower in a low prevalence setting, e.g. when testing the asymptomatic population. Another limitation of this validation study lies in the limited sample size leading to broad 95% confidence intervals, requiring confirmation of these data at a larger scale. Also, here we used plasma and the test was performed in a laboratory environment; we may expect different results in real-life at patients’ bed and using capillary blood. Finally, our present conclusions only apply to the Augurix RDT, and must not be generalized to other currently available RDTs.

In conclusion, Augurix RDT is not meant to replace a SARS-CoV-2 RT-PCR diagnostic test in the first week of the disease, but could be a reliable option for assessing the SARS-CoV-2 serology in moderate to high COVID-19 prevalence settings, i.e. when testing the sub-population of individuals having presented COVID-19 symptoms, especially in situations where automated ECLIA or ELISA are not available, with samples collected between at least 15 days and up to 180 days after the onset of symptoms.

## Ethical consideration

This study was evaluated by the Ethics Committee of Canton de Vaud, Lausanne, Switzerland (CER-VD) and they judged that it did not deserve a specific approval being only a quality assessment of diagnostic tests with foreign residual samples.

All necessary patient/participant consent has been obtained by the supplier of residual samples and the appropriate institutional forms have been archived.

## Data Availability

Data are available in Supplementary Material

## Acknowledgments

We thank Augurix SA (Monthey, Switzerland) for providing Simtomax CoronaCheck RDTs free-of-charge.

## Funding

This research was financially supported by the HES-SO Valais-Wallis.

## Conflict of interest

Percevent J Ducrest has a R&D mandate with Augurix SA, the manufacturer of the RDT used in this study. Augurix SA had no role in the study design, the realization nor in result interpretation. The other authors have no conflict of interest to disclose.

## Contributions

**Percevent J Ducrest:** Conceptualization, Methodology, Investigation, Formal analysis, Writing - Original Draft preparation. **Antoine Freymond:** Investigation, Formal analysis, Writing - Original Draft preparation. **Jean-Manuel Segura:** Writing - Original Draft preparation, Writing - Review & Editing preparation, Visualization

