## Supplementary Material for "Performance evaluation of the Simtomax^®^ CoronaCheck rapid diagnostic test"

#### RT-PCR Positive samples

| Sample # | Age Range (gender) | DPS | IgG | IgM | Elecsys® Anti-SARS-CoV-2 total Ig (COI) |
|---|---|---|---|---|---|
| 1 | >50 (F) | 180 | POS | NEG | NT |
| 2 | >50 (M) | 170 | POS | NEG | NT |
| 3 | <20 (F) | 57 | POS | NEG | NT |
| 4 | >50 (F) | 15 | POS | NEG | NT |
| 5 | >50 (F) | 22 | POS | POS | NT |
| 6 | >50 (F) | 14 | POS | NEG | NT |
| 7 | 30-40 (M) | 17 | POS | NEG | NT |
| 8 | >50 (M) | 19 | POS | NEG | NT |
| 9 | 30-40 (M) | 19 | POS | NEG | NT |
| 10 | >50 (F) | 17 | POS | NEG | NT |
| 11 | 20-30 (F) | 20 | POS | NEG | NT |
| 12 | >50 (F) | 21 | POS | NEG | NT |
| 13 | >50 (M) | 11 | POS | NEG | NT |
| 14 | 30-40 (F) | 36 | POS | POS | NT |
| 15 | 40-50 (M) | 19 | POS | NEG | NT |
| 16 | 20-30 (F) | 16 | **NEG** | **NEG** | NT |
| 17 | 20-30 (F) | 30 | POS | NEG | NT |
| 18 | 30-40 (F) | 24 | POS | NEG | NT |
| 19 | >50 (M) | 14 | **NEG** | **NEG** | NT |
| 20 | >50 (F) | 27 | POS | POS | NT |
| 21 | 40-50 (F) | 21 | POS | NEG | NT |
| 22 | 20-30 (F) | 22 | POS | POS | NT |
| 23 | >50 (M) | 18 | POS | NEG | NT |
| 24 | >50 (M) | 16 | POS | NEG | NT |
| 25 | >50 (M) | 36 | POS | POS | NT |
| 26 | >50 (M) | 39 | POS | POS | 180,9 |
| 27 | <20 (M) | 22 | POS | NEG | 169,6 |
| 28 | 20-30 (F) | 35 | POS | POS | 154,2 |
| 29 | <20 (M) | 34 | POS | NEG | 130,8 |
| 30 | >50 (F) | 27 | POS | NEG | 126,7 |
| 31 | <20 (M) | 20 | POS | POS | 114,9 |
| 32 | 30-40 (M) | 41 | POS | POS | 107,7 |
| 33 | >50 (M) | 14 | POS | POS | 91,45 |
| 34 | 30-40 (M) | 28 | POS | NEG | 89,2 |
| 35 | >50 (M) | 33 | POS | POS | 85,5 |
| 36 | <20 (F) | 41 | POS | POS | 76,85 |
| 37 | <20 (M) | 27 | POS | NEG | 63,4 |
| 38 | 30-40 (F) | 32 | POS | NEG | 58,6 |
| 39 | >50 (M) | 10 | POS | POS | 51,1 |
| 40 | >50 (M) | 16 | NEG | POS | 49,05 |

| | | | | | |
|---|---|---|---|---|---|
| 41 | 40-50 (M) | 20 | POS | POS | 42,2 |
| 42 | >50 (F) | 19 | POS | POS | 36,9 |
| 43 | >50 (F) | 17 | POS | NEG | 20,16 |
| 44 | 30-40 (F) | 24 | POS | NEG | 10,1 |
| 45 | >50 (F) | 15 | NEG | POS | 4,98 |
| 46 | >50 (M) | 26 | POS | NEG | 2,06 |
| 47 | 40-50 (M) | 13 | **NEG** | **NEG** | 1,43 |
| 48 | 30-40 (F) | 10 | **NEG** | **NEG** | 1,19 |

NT: not tested

### Control Group Samples

| Sample # | Age (gender) | IgG | IgM |
|---|---|---|---|
| 1 | 20-30 (M) | NEG | NEG |
| 2 | 20-30 (M) | NEG | NEG |
| 3 | >50 (M) | NEG | NEG |
| 4 | 40-50 (F) | NEG | NEG |
| 5 | >50 (F) | NEG | NEG |
| 6 | 20-30 (F) | NEG | POS |
| 7 | 30-40 (M) | NEG | NEG |
| 8 | >50 (M) | NEG | NEG |
| 9 | 20-30 (M) | NEG | NEG |
| 10 | 40-50 (M) | NEG | NEG |
| 11 | 40-50 (F) | NEG | NEG |
| 12 | 20-30 (M) | NEG | NEG |
| 13 | 20-30 (F) | NEG | NEG |
| 14 | 30-40 (F) | NEG | NEG |
| 15 | 20-30 (M) | NEG | NEG |
| 16 | >50 (F) | NEG | NEG |
| 17 | >50 (F) | NEG | NEG |
| 18 | 40-50 (F) | NEG | NEG |
| 19 | 30-40 (M) | NEG | NEG |
| 20 | 40-50 (F) | NEG | NEG |
| 21 | 30-40 (F) | NEG | NEG |
| 22 | >50 (M) | NEG | NEG |
| 23 | 30-40 (M) | NEG | NEG |
| 24 | 20-30 (M) | NEG | NEG |
| 25 | 40-50 (M) | NEG | NEG |
| 26 | >50 (F) | NEG | NEG |
| 27 | 20-30 (F) | NEG | NEG |
| 28 | 20-30 (F) | NEG | NEG |
| 29 | 30-40 (M) | NEG | NEG |
| 30 | 30-40 (F) | NEG | NEG |
| 31 | 30-40 (M) | NEG | NEG |
| 32 | 30-40 (M) | NEG | NEG |
| 33 | 30-40 (F) | NEG | NEG |
| 34 | 40-50 (M) | NEG | NEG |
| 35 | 30-40 (F) | NEG | NEG |
| 36 | 30-40 (M) | NEG | NEG |
| 37 | 20-30 (F) | NEG | NEG |

| | | | |
|---|---|---|---|
| 38 | 20-30 (F) | NEG | NEG |
| 39 | 30-40 (M) | NEG | NEG |
| 40 | >50 (M) | NEG | NEG |
| 41 | >50 (F) | NEG | NEG |
| 42 | >50 (M) | NEG | NEG |
| 43 | 20-30 (M) | NEG | NEG |
| 44 | >50 (M) | NEG | NEG |
| 45 | 30-40 (M) | NEG | NEG |
| 46 | >50 (M) | NEG | NEG |
| 47 | >50 (F) | NEG | NEG |
| 48 | >50 (F) | NEG | NEG |
| 49 | 30-40 (M) | NEG | NEG |
| 50 | 20-30 (F) | NEG | NEG |
| 51 | 30-40 (F) | NEG | NEG |
| 52 | 40-50 (F) | NEG | NEG |
| 53 | 30-40 (F) | NEG | NEG |
| 54 | 40-50 (F) | NEG | NEG |
| 55 | 30-40 (M) | NEG | NEG |
| 56 | >50 (M) | NEG | NEG |
| 57 | 40-50 (F) | NEG | NEG |
| 58 | >50 (M) | NEG | POS |
| 59 | 20-30 (M) | NEG | NEG |
| 60 | 30-40 (F) | NEG | NEG |
| 61 | 30-40 (M) | NEG | NEG |
| 62 | 40-50 (M) | NEG | NEG |
| 63 | >50 (M) | NEG | NEG |
| 64 | 30-40 (F) | NEG | NEG |
| 65 | 30-40 (F) | NEG | NEG |
| 66 | >50 (F) | NEG | NEG |
| 67 | 40-50 (M) | NEG | NEG |
| 68 | <20 (F) | NEG | NEG |
| 69 | 20-30 (F) | NEG | NEG |
| 70 | 30-40 (F) | NEG | NEG |
| 71 | 30-40 (F) | NEG | NEG |
| 72 | 30-40 (M) | NEG | NEG |
| 73 | >50 (F) | NEG | NEG |
| 74 | 30-40 (F) | NEG | NEG |
| 75 | <20 (M) | NEG | NEG |
| 76 | >50 (F) | NEG | NEG |
| 77 | 40-50 (M) | NEG | NEG |
| 78 | 40-50 (F) | NEG | NEG |
| 79 | 30-40 (M) | NEG | NEG |
| 80 | >50 (F) | NEG | NEG |
| 81 | 20-30 (M) | NEG | NEG |
| 82 | >50 (F) | NEG | NEG |
| 83 | 20-30 (F) | NEG | NEG |
| 84 | 40-50 (F) | NEG | POS |
| 85 | >50 (F) | NEG | NEG |

| | | | |
|---|---|---|---|
| 86 | 40-50 (M) | NEG | NEG |
| 87 | 40-50 (M) | NEG | NEG |
| 88 | >50 (M) | NEG | NEG |
| 89 | 30-40 (F) | NEG | NEG |
| 90 | 20-30 (F) | NEG | NEG |
| 91 | >50 (M) | NEG | NEG |
| 92 | 30-40 (M) | NEG | NEG |
| 93 | >50 (F) | NEG | NEG |
| 94 | 40-50 (F) | NEG | NEG |
| 95 | 40-50 (F) | NEG | NEG |
| 96 | >50 (M) | NEG | NEG |
| 97 | >50 (M) | NEG | NEG |
| 98 | 20-30 (F) | NEG | NEG |

na: unknown
